# Escape from the Past: Improved Risk Stratification in Advanced Heart Failure using Novel Hemodynamic Parameters

**DOI:** 10.1101/2023.04.18.23288774

**Authors:** the FLIGHT Working Group, Nicole Cyrille-Superville, Sriram D Rao, Jason P Feliberti, Priyesh A Patel, Kamala Swayampakala, Shashank S Sinha, Eric I Jeng, Rohan M Goswami, David F Snipelisky, Adam D DeVore, Samer S Najjar, Jonathan Grinstein

**Author notes:** Corresponding Author: Nicole Cyrille-Superville, MD, Advanced Heart Failure, Sanger Heart and Vascular Institute, Atrium Health, 1237 Harding Place, Suite 4100, Charlotte NC 28204.

## Abstract

**Objectives:** We sought to determine whether novel hemodynamic parameters provide added prognostic value in a real world heart failure population undergoing right heart catheterization (RHC) for consideration of advanced surgical therapies.

**Background:** Invasive hemodynamics are fundamental in the assessment of patients with advanced heart failure. Early studies showed that right atrial pressure (RAP) and pulmonary capillary wedge pressure (PCWP), were significantly associated with long term outcomes, but cardiac index (CI) was not. In recent years, there has been a number of novel hemodynamic parameters such as cardiac power output (CPO), pulmonary artery pulsatility index (PAPI), and aortic pulsatility index (API) however it remains unclear, in part due to variability in study population and design, which hemodynamic parameters are most prognostic in regard to long-term need for OHT or left ventricular assist device (LVAD).

**Methods:** This study is a retrospective review of patients from the PRognostic Evaluation During Invasive CaTheterization for Heart Failure (PREDICT-HF) registry. The demographics, laboratory variables, vital signs, hemodynamic variables and outcomes of patients who underwent RHC at one of the 9 member institutions were collected. The cumulative endpoint for this analysis was survival to OHT or LVAD, or death within 6 months of RHC.

**Results:** A total of 846 patients were included for the analysis, of which 176 (21%) met the primary endpoint. The majority of those within the primary outcome either underwent LVAD implant (n=76, 42%) or died (n=75, 42%). On multivariate analysis, a model incorporating the traditional hemodynamic variables, PCWP (OR 1.10, 10.4-1.15, p < 0.001) and CI (OR 0.86, 0.81-0.92, p<0.001) were shown to be predictive of adverse outcomes. In a separate multivariate model incorporating novel variables, CPO (OR 0.76, 0.71-0.83, p<0.001), API (OR 0.94, 0.91-0.96, P < 0.001) and PAPI (OR 1.02, 1.00-1.03, p 0.027) were all predictive. Moreover, patients with positively concordant API and CPO had the best freedom from endpoint (94.7%), whilst those with negatively concordant API and CPO had the worst freedom from endpoint (61.5%, p < 0.001). Those with discordant API and CPO had similar freedom from endpoint (high API and low CPO 83.7%; low API and high CPO 89.7%).

**Conclusion:** The novel hemodynamic parameters of API and CPO are highly predictive of the need for OHT or LVAD or death within 6 months and in combination offer added prognostic value.

## Introduction

Heart failure (HF) in the United States is a serious public health concern affecting over 6.5 million patients and 650, 000 new cases diagnosed annually ^1^. Despite progress in treatment, an estimated 1-10% of patients progress to advanced heart failure (AHF) with dismal 5 year survival rates among Stage C and D patients of 75% and 20% respectively ^2,3^. Thus, accurate prognostication is of great importance to allow timely referral for consideration of advanced therapies, namely orthotopic heart transplantation (OHT) and durable mechanical circulatory support (MCS).

Invasive hemodynamics are fundamental to characterization and prognostication in AHF. Nevertheless, it’s unclear which hemodynamic parameters are most prognostic particularly with regard to long-term outcomes such as need for advanced therapies. Moreover, an important consideration with the advent of hemodynamic parameters such as cardiac power output (CPO), pulmonary artery pulsatility index (PAPI), and aortic pulsatility index (API) is whether these novel parameters in isolation or in combination can be used for long term prognostication.

An exploratory analysis of the ESCAPE trial, for example, showed that filling pressures, including right atrial pressure (RAP) and pulmonary capillary wedge pressure (PCWP), were significantly associated with the primary outcome of combined risk of death, cardiovascular hospitalization and OHT, but cardiac index (CI) was not ^4,5^. Since then, numerous novel indices have been reported to show prognostic value. For example, PAPI, calculated as the ratio of pulmonary artery pulse pressure to right atrial pressure, less than 1.7 was found to be a significant predictor of death or hospitalization at 6 months ^6^. Subsequently, Belkin et found that the API, calculated as (systolic-diastolic blood pressure)/PCWP) >2.9 was associated with decreased risk of death and need for left ventricular assist device (LVAD) or OHT at 6 months^7^. Cardiac power output (CPO) has been shown to correlate strongly with in-hospital mortality in a patient population of cardiogenic shock secondary to acute myocardial infarction (AMI) but did not show a correlation with risk of death/LVAD/OHT at 6 months in a less acutely ill patient population ^7,8^.

Application of various hemodynamic variables and risk scores for prognostication is challenging in practice due to substantial variability in trial design and study cohorts, since most data were derived and validated in selected clinical trial populations or single center studies. Here, we report that novel hemodynamic parameters provide important prognostic information in a “real-world” US-based cohort of heart failure patients across several centers spanning large geographic range.

## Methods

The PRognostic Evaluation During Invasive CaTheterization for Heart Failure (PREDICT-HF) is a registry comprised of retrospective patient data from 10 member institutions of the Southeast Future Leaders in Growing Heart Failure Therapies (SE-FLIGHT) program. (Supplemental Material) The study was approved by the central IRB at Atrium Heath (IRB #02-21-06E) with independent IRB approval and data sharing agreements subsequently obtained for all participating sites. Atrium Health served as the data coordinating center and performed all statistical analysis. Charts were reviewed from January 1^st^ 2013 to December 31^st^ 2019 and patients with chronic or acute on chronic HF undergoing isolated right heart catheterization (RHC) were included in the analysis. Patients with AMI, those undergoing concomitant left heart catheterization for revascularization or valvular procedures such as mitral clip or transcatheter aortic valve replacement at the time of RHC were excluded. Those undergoing RHC solely for the assessment of pulmonary hypertension were also excluded. A full list of inclusion and exclusion criteria is included in the Supplemental Material. Patient demographics, laboratory variables, vital signs and hemodynamic variables were collected. The cumulative endpoint for this analysis was survival to surgical advanced heart failure therapy (LVAD implantation or OHT) or death within 6 months of RHC.

### Statistical analysis

Demographics, past medical history, baseline lab and hemodynamic values were summarized as frequencies and percentages for categorical variables and as means (± standard deviation) for continuous variables and compared between patients that received medical management versus advanced therapies or death with either Student t tests or Mann-Whitney U (Wilcoxon) tests depending on normality as determined by Shapiro-Wilk tests for continuous variables, and chi-square or Fisher’s exact tests for categorical variables. Relationships between baseline characteristics, lab and hemodynamic variables were tested using univariate logistic regression analysis. Covariates with a p-value of <0.05 on univariate regression analysis or based on previously established clinical relevance were selected into the multivariable logistic regression models and results were presented as odds ratio (OR) and 95% confidence interval. Hemodynamic variables were checked for multicollinearity using Spearman rank correlations and two separate adjusted models were run so there were no multicollinearity issues between the traditional and more modern hemodynamic variables. Receiver operator characteristic (ROC) curves were used to determine the appropriate cutoff values, as well as the sensitivity, specificity, correctly classified, and area under the curve (AUC) values for each of the six hemodynamic variables. Kaplan–Meier time-to-event analysis was conducted to describe time to the composite endpoint, and then tested using log rank tests. To assess whether the combination of API and CPO were more discriminatory for the outcome of interest, we compared logistic regression models with CPO and API cutpoints and discriminant ability was assessed with concordance (c) statistics. All tests were two-tailed and considered statistically significant with a p-value <0.05. All statistical analyses were performed using SAS Enterprise Guide 7.1 (SAS Institute, Cary, North Carolina).

## Results

A total of 846 patients were included for analysis. The average age was 58.8 years old with 43% female and 50.2% African American. Baseline demographics are presented in Table 1. Of the 846 patients included, 176 (21%) met the primary endpoint. Patients of male gender or a past medical history of ventricular tachycardia were more likely to meet the primary endpoint. Laboratory values at baseline showed White Blood Cell, Sodium, Blood Urea Nitrogen, Creatinine and Alanine Aminotransferase were all significantly different between the two cohorts. The majority of those who met the primary outcome either underwent LVAD implantation (n=76, 42%) or died (n=75, 42%) compared to underwent transplant (n=28, 16%).

**Table 1.** Baseline Demographic Characteristics, Laboratory and Right Heart Catheterization measurements of Study participants.

|  | Study Group<br>(N=846) |  | Transplant or<br>LVAD or<br>Death in 6-months<br>(n=176 (21%)) |  | Only Medical<br>Management<br>(n=667 (79%)) |  | p-value |
| --- | --- | --- | --- | --- | --- | --- | --- |
|  | n | % | n | % | n | % |  |
| Baseline Characteristics |  |  |  |  |  |  |  |
| Age (Mean ±SD) | 58.8 | 14.5 | 58.8 | 14.5 | 59.20 | 13.6 | 0.7735 |
| Female | 366 | 43.3% | 61 | 34.1% | 305 | 45.7% | 0.0050 |
| Race |  |  |  |  |  |  | 0.1843 |
| White | 331 | 39.1% | 80 | 44.7% | 251 | 37.6% |  |
| African American | 425 | 50.2% | 84 | 46.9% | 341 | 51.1% |  |
| Other | 90 | 10.6% | 15 | 8.4% | 75 | 11.2% |  |
| Past Medical History |  |  |  |  |  |  |  |
| Non-Ischemic Cardiomyopathy | 201 | 23.8% | 56 | 31.3% | 145 | 21.7% | 0.1165 |
| Coronary Artery Disease | 329 | 38.9% | 74 | 41.3% | 255 | 38.2% | 0.4485 |
| Hyperlipidemia | 267 | 31.6% | 67 | 37.4% | 200 | 30.0% | 0.0570 |
| Peripheral Vascular Disease | 43 | 5.1% | 11 | 6.1% | 32 | 4.8% | 0.4661 |
| Obstructive Sleep Apnea | 172 | 20.3% | 45 | 25.1% | 127 | 19.0% | 0.0718 |
| Atrial fibrillation | 157 | 18.6% | 32 | 17.9% | 125 | 18.7% | 0.7919 |
| Ventricular Tachycardia | 88 | 10.4% | 29 | 16.2% | 59 | 8.8% | 0.0042 |
| Baseline Laboratory values (Mean ±SD) |  |  |  |  |  |  |  |
| White Blood Cell | 8.0 | 4.0 | 8.4 | 5.0 | 7.9 | 3.6 | 0.2099 |
| Hemoglobin | 12.3 | 2.3 | 12.3 | 2.2 | 12.2 | 2.3 | 0.6727 |
| Platelets | 223.3 | 88.3 | 206.6 | 88.8 | 228.6 | 87.5 | 0.0056 |
| Sodium | 138.9 | 4.1 | 138.1 | 4.7 | 139.2 | 3.9 | 0.0114 |
| Potassium | 4.2 | 0.6 | 4.2 | 0.6 | 4.2 | 0.6 | 0.6684 |
| Blood Urea Nitrogen | 30.0 | 20.4 | 35.6 | 25.1 | 28.2 | 18.4 | <0.0001 |
| Creatinine | 1.7 | 1.5 | 1.9 | 1.8 | 1.6 | 1.3 | 0.023 |
| Albumin | 3.6 | 0.6 | 3.5 | 0.6 | 3.7 | 0.6 | 0.0688 |
| Alanine Aminotransferase U/L | 57.2 | 209.8 | 105.0 | 373.5 | 40.8 | 102.3 | 0.0014 |
| Aspartate Aminotransferase U/L | 65.4 | 303.8 | 128.4 | 562.4 | 43.6 | 116.2 | 0.0717 |
| Baseline Hemodynamics (Mean ±SD) |  |  |  |  |  |  |  |
| Right atrial pressure (mmHg) | 11.0 | 6.6 | 13.0 | 7.6 | 10.5 | 6.2 | <0.0001 |
| Systolic blood pressure | 122.0 | 21.9 | 113.7 | 16.5 | 124.2 | 22.6 | <0.0001 |
| Diastolic blood pressure | 72.8 | 13.0 | 71.3 | 12.5 | 73.2 | 13.1 | 0.074 |
| Mean Arterial Pressure | 89.2 | 14.0 | 85.4 | 12.2 | 90.2 | 14.3 | <0.0001 |
| Mean pulmonary arterial pressure (mmHg) | 31.7 | 11.6 | 36.2 | 10.6 | 30.5 | 11.6 | <0.0001 |
| Pulmonary artery systolic pressure (mmHg) | 47.1 | 17.1 | 52.6 | 15.4 | 45.6 | 17.3 | <0.0001 |
| Pulmonary artery diastolic pressure (mmHg) | 22.6 | 9.2 | 26.1 | 8.5 | 21.7 | 9.2 | <0.0001 |
| Pulmonary capillary wedge pressure (mmHg) | 20.0 | 9.4 | 24.6 | 9.6 | 18.8 | 9.0 | <0.0001 |
| Pulmonary artery pulsatility index | 3.2 | 2.9 | 3.0 | 2.7 | 3.2 | 2.9 | <0.0001 |
| Aortic pulsatility Index | 3.2 | 2.3 | 2.1 | 1.5 | 3.5 | 2.4 | <0.0001 |
| Cardiac Power Output | 1.0 | 0.5 | 0.8 | 0.3 | 1.1 | 0.5 | <0.0001 |
| FICK Cardiac Output (L/min) | 5.1 | 2.2 | 4.1 | 1.5 | 5.4 | 2.3 | <0.0001 |
| FICK Cardiac Index (L/min/m<sup>2</sup>) | 2.5 | 1.3 | 2.1 | 1.3 | 2.7 | 1.3 | **<0.0001** |

### Hemodynamic Predictors of surgical heart failure therapies or death

We then assessed the effects of individual RHC measurements and the primary outcome. As seen in Table 1, RAP, systolic blood pressure (SBP), mean arterial pressure (MAP), mean pulmonary artery pressure (mPAP). Pulmonary artery systolic pressure (PASP), pulmonary artery diastolic pressure (PADP), PCWP, API, PAPI, CPO, Fick CO and Fick CI were significantly different between the two groups. On a Univariate analysis RAP, PCWP, API, CPO and Fick CI were all shown to be correlated with the primary outcome (Figure 1). Results from multivariable models assessing the association between traditional and more modern hemodynamic variables with outcome of interest are presented in Table 2. In the model incorporating the traditional variables, PCWP (OR:1.10, 95% CI: 1.04-1.15, p < 0.001) and CI (OR:0.86, 95% CI:0.81-0.92, p<0.001) were showed to be predictive of adverse outcomes. In the model incorporating novel variables, all 3 variables were shown to be significant, with CPO (OR:0.76, 95% CI:0.71-0.83, p<0.001) and API (OR:0.94, 95% CI:0.91-0.96, P < 0.001) having more robust association than PAPI (OR:1.02, 95% CI:1.00-1.03, p 0.027).

**Table 2.** Multivariate Analyses of Hemodynamic Variables.

|  | **OR** | **95% CI** |  | **p-value** |
| --- | --- | --- | --- | --- |
| **Multivariable Analysis Model-I** |  |  |  |  |
| API (0.2 units increase) | 0.94 | 0.91 | 0.96 | **<0.001** |
| PAPI (0.2 units increase) | 1.02 | 1.00 | 1.03 | **0.0267** |
| CPO (0.1 units increase) | 0.76 | 0.71 | 0.83 | **<0.001** |
| **Multivariable Analysis Model-II** |  |  |  |  |
| PCWP (2 units increase) | 1.10 | 1.04 | 1.15 | **<0.001** |
| RAP (2 units increase) | 0.99 | 0.93 | 1.07 | 0.8551 |
| FICK CI (0.2 units increase) | 0.86 | 0.81 | 0.92 | **<0.001** |

**Figure 1.**
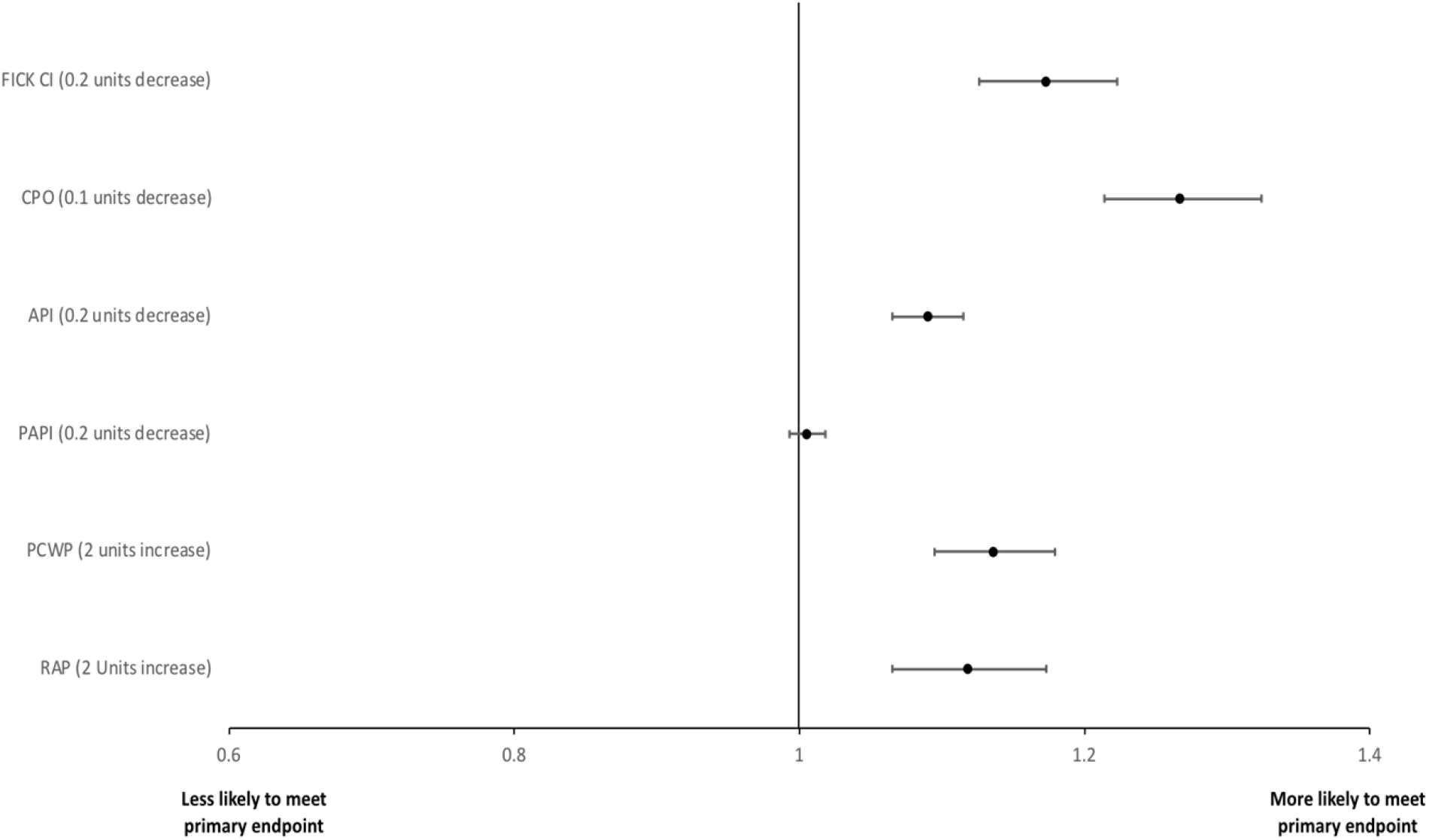
Univariate analysis of hemodynamic Parameters.

### Cutoff derivation and Predictive value of hemodynamic variables

We subsequently used ROC analysis to create cutoffs for each of the variables that were shown to be significant in our multivariable model (Supplemental analysis). Kaplan Meier survival analysis showed that CPO (cutoff 0.8, p < 0.001), API (cutoff 2.3, p < 0.001), PAPI (cutoff 1.3, p < 0.016), RAP (cutoff 14mmHg, p < 0.001), PCWP (cutoff 21mmHg, p < 0.001) and Fick CI (cutoff 2.3, p < 0.001) were all predictive of the primary outcome (Figure 2).

**Figure 2.**
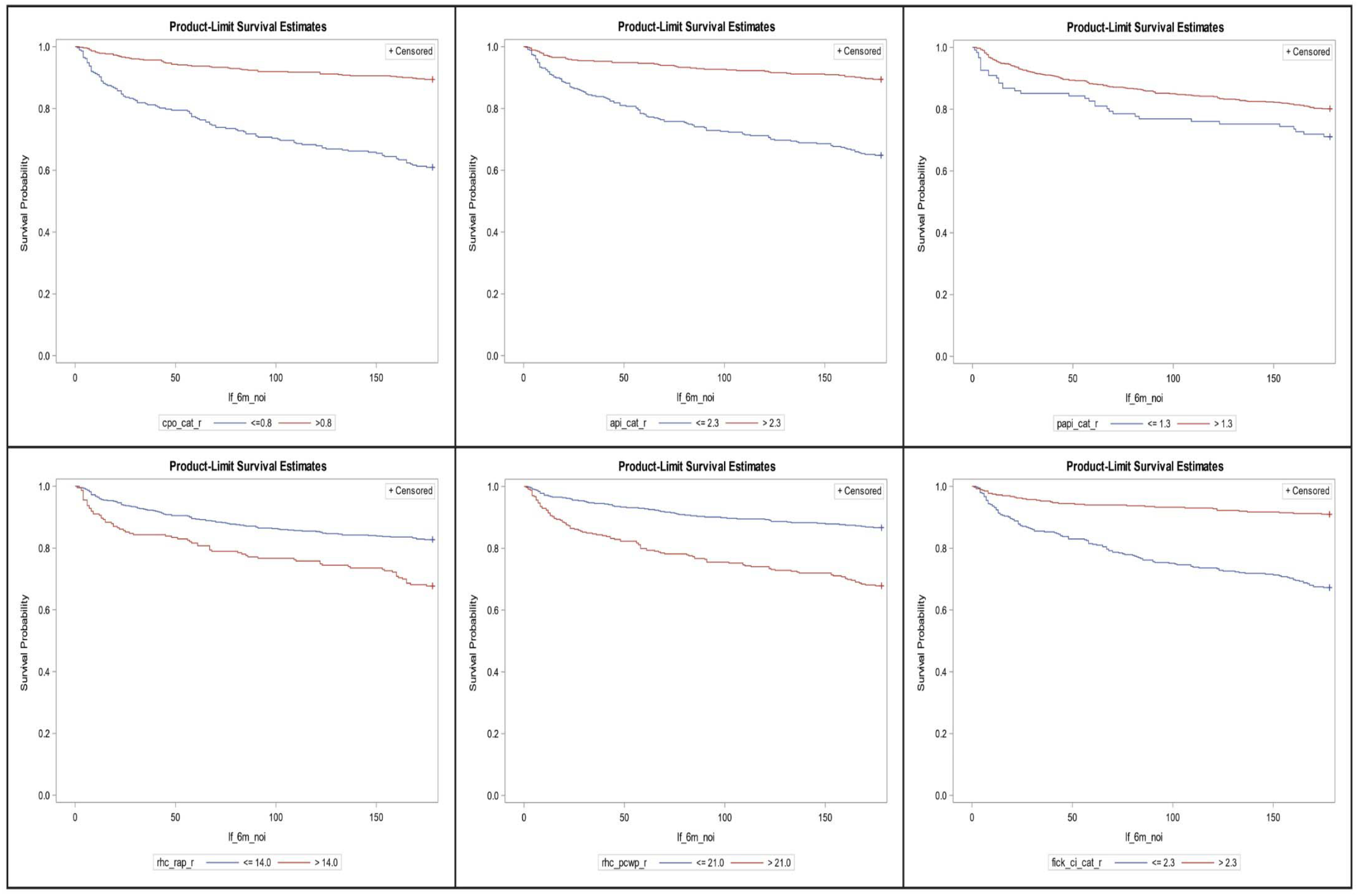
Kaplan Meier Curves showing the Predictive Value of Hemodynamic Variables.

When API and CPO were used in combination, patients with positively concordant API and CPO had the best freedom from endpoint (94.7%), whilst those with negatively concordant API and CPO had the worst freedom from endpoint (61.5%, p < 0.001). Those with discordant API and CPO had similar freedom from endpoint (high API and low CPO 83.7%; low API and high CPO 89.7%). A model with API and CPO together (Figure 3. Panel A) performed better in predicting advanced treatments or death within 6-months with a c-statistic of 0.75 (95% CI: 0.71-0.79) than a model with either API (c-statistic 0.68, 95% CI: 0.64-0.72) or CPO (c-statistic 0.70, 95% CI: 0.66-0.74) alone.

**Figure 3.**
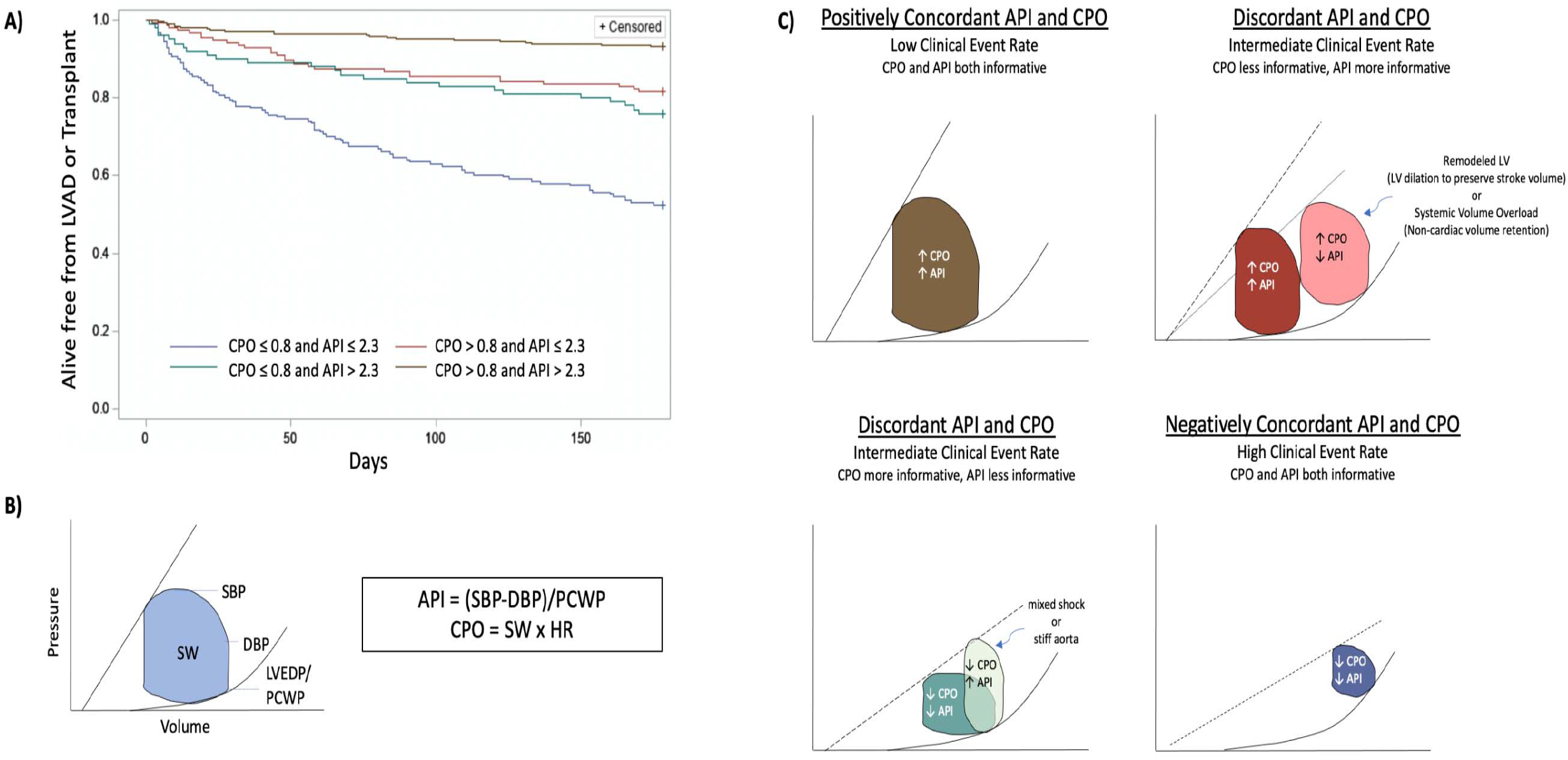
1. Improved Risk Stratification of Advanced Heart Failure using novel hemodynamic parameters, API and CPO 2. API and CPO measurements depicted utilizing pressure-volume loops 3. Pressure-volume loops demonstrating the relationship and utility of API and CPO in different clinical states

## Discussion

In this current, multi-institutional analysis of all-comers with systolic heart failure who underwent RHC, advanced hemodynamic metrics were found to be highly prognostic at predicting 6-month need for advanced therapies or death. The predominant findings from our analysis are as follows: 1) The advanced hemodynamic parameters have an added prognostic role above established clinical parameters, 2) API and CPO are highly prognostic of the need for advanced surgical therapies or death at 6 months and 3) There is additive value for risk stratification using API and CPO concomitantly when compared to API or CPO alone.

Identification of the prognostic potential of both standard and advanced hemodynamic variables has been historically challenging and often times confusing ^9-11^. Cardiac index, once thought to be the quintessential hemodynamic parameter, has shown mixed results for prognostication ^9,11,12^. Even in the more contemporaneous era, CI continues to have mixed results. In a sub-analysis of ESCAPE trial, residual congestion and not CI was predictive of 6-month events ^5^. In a robust registry from the Veteran Affairs Administration, Thermodilution CI was more predictive of mortality than estimated Fick CI with poor agreement between the two measurements ^13^. Conversely, the congestive state of the patient has shown to be a more reliable prognostic marker ^9,11,12,14^. Both an elevated RAP and PCWP portend a poor outcome and need for advanced therapies or death ^5^. CPO similarly has shown mixed results in terms of prognostication. In the shock trial, CPO was found to be the strongest predictor of mortality, however more recently CPO failed to discriminate clinical events in the Cardiogenic Shock Working Group analysis (CSWG) ^8,15,16^.

The heterogeneity of the hemodynamic parameters is largely driven by variable patient populations and patient acuity as well as different eras of clinical medicine including background device and medication optimization. When patients are in cardiogenic shock, it is often straightforward to identify patient acuity, however, this task is often challenging in ambulatory patients with chronic HF. Hemodynamic parameters may have different relative weights depending on the stage and severity of disease as well as the mechanism of shock. For example, the Shock trial, which showed CPO to be highly prognostic, was conducted in a patient population of acute MI shock whereas the CSWG analysis was conducted in a patient population of cardiogenic shock related to both AMI and chronic HF ^8,16^.

We postulate that the chronicity of shock and the patient’s physiologic response may influence the discriminatory potential of some of the hemodynamic parameters. In response to an acute drop in contractility, such as following a myocardial infarction, the end systolic pressure volume relationship shifts downward (reduction in end-systolic elastance Ees) leading to an immediate reduction in stroke volume, stroke work (SW), cardiac output and CPO (Figure 3. Panel C). If the body has a chance to remodel, activation of the renin-angiotensin-aldosterone axis leads to retention of salt and water leading to a rightward shift along the end-diastolic pressure volume relationship ^17^. Depending on the afterload and the contractile state of the patient, this often normalizes stroke volume, SW, CO and CPO. Under these conditions, CPO may not fully reflect the clinical state of the patient whereas API would still be prognostic. Conversely, API can be influenced by the aortic properties and vascular resistance which can influence the pulse pressure (PP) independent from cardiac output. Thus, in settings of mixed shock or in patients with stiff aortic vasculature, API may be high and not fully reflect the clinical state whereas CPO would be more prognostic in this context. The simultaneous use of API and CPO can overcome the individual limitations of each parameter in isolation. A negative concordant low API and CPO reflects a rightward and downward shifted pressure volume loop and reflects a patient who is both congested and in a low output state (Figure 3. Panel C).

The current heart failure guidelines have downplayed the role of continuous pulmonary artery pressure monitoring in routine patient care ^18^. The guideline recommendations largely reflect the disparate literature surrounding the role of hemodynamic monitoring in the day-to-day care of a hospitalized patient. A complete hemodynamic assessment still has an important role in prognostication and suitability of advanced therapies. Here we show that API and CPO, both in isolation and in combination, have an important role in assessing the need for advanced therapies. The current UNOS allocation system heavily relies on CI, SBP and PCWP to predict waitlist mortality. Additional studies are needed to determine if the addition of advanced hemodynamic parameters can improve risk assessment at the time or transplant as it relates to status designation at the time of listing ^19^.

### Limitations

There are several limitations in this analysis. First, this is a retrospective cohort study, and as such there are inherent limitations and residual confounding that cannot be excluded with this type of analysis. Second, there was a non-standard distribution of patients in the cohort from the different participating centers, both individually and geographically. When reviewing center level data, there was a lower clinical event rate in the largest contributing center, and as such any bias would be towards the null hypothesis which, in turn, strengthens the validity of the results. Third, the inclusion of a number of centers with differing practice patterns may have effects on patient management, especially in regard to delivery of advanced therapeutic options. The variability in practice patterns is even further exacerbated by the inclusion period straddling the changes to the UNOS allocation system in 2018. We sought to account for this by our use of the composite endpoint that incorporated both transplant and LVAD as well as mortality but validation in other data sources will be important prior to incorporation of this into clinical guidelines.

## Conclusions

A complete hemodynamic assessment can aid in the prognostication of AHF. Specifically, the advanced hemodynamic parameters of API and CPO were highly predictive of the need for advanced surgical therapies or death, and that when used in combination they had added predictive value. API and CPO should be incorporated into routine risk stratification.

## Data Availability

All data analyzed and generated is available at Atrium Health in a secured Redcap database and is available upon request.

## Acknowledgements

All listed authors contributed to the manuscript.

## References

1. Go AS, Mozaffarian D, Roger VL, Benjamin EJ, Berry JD, Borden WB, Bravata DM, Dai S, Ford ES, Fox CS, et al. Heart disease and stroke statistics--2013 update: a report from the American Heart Association. Circulation. 2013;127:e6–e245. doi: 10.1161/CIR.0b013e31828124ad

2. Crespo-Leiro MG, Metra M, Lund LH, Milicic D, Costanzo MR, Filippatos G, Gustafsson F, Tsui S, Barge-Caballero E, De Jonge N, et al. Advanced heart failure: a position statement of the Heart Failure Association of the European Society of Cardiology. Eur J Heart Fail. 2018;20:1505–1535. doi: 10.1002/ejhf.1236

3. Ammar KA, Jacobsen SJ, Mahoney DW, Kors JA, Redfield MM, Burnett JC, Rodeheffer RJ. Prevalence and prognostic significance of heart failure stages: application of the American College of Cardiology/American Heart Association heart failure staging criteria in the community. Circulation. 2007;115:1563–1570. doi: 10.1161/CIRCULATIONAHA.106.666818

4. Binanay C, Califf RM, Hasselblad V, O’Connor CM, Shah MR, Sopko G, Stevenson LW, Francis GS, Leier CV, Miller LW, et al. Evaluation study of congestive heart failure and pulmonary artery catheterization effectiveness: the ESCAPE trial. JAMA. 2005;294:1625–1633. doi: 10.1001/jama.294.13.1625

5. Cooper LB, Mentz RJ, Stevens SR, Felker GM, Lombardi C, Metra M, Stevenson LW, O’Connor CM, Milano CA, Patel CB, et al. Hemodynamic Predictors of Heart Failure Morbidity and Mortality: Fluid or Flow? J Card Fail. 2016;22:182–189. doi: 10.1016/j.cardfail.2015.11.012

6. Kochav SM, Flores RJ, Truby LK, Topkara VK. Prognostic Impact of Pulmonary Artery Pulsatility Index (PAPi) in Patients With Advanced Heart Failure: Insights From the ESCAPE Trial. J Card Fail. 2018;24:453–459. doi: 10.1016/j.cardfail.2018.03.008

7. Belkin MN, Alenghat FJ, Besser SA, Nguyen AB, Chung BB, Smith BA, Kalantari S, Sarswat N, Blair JEA, Kim GH, et al. Aortic pulsatility index predicts clinical outcomes in heart failure: a sub-analysis of the ESCAPE trial. ESC Heart Fail. 2021;8:1522–1530. doi: 10.1002/ehf2.13246

8. Fincke R, Hochman JS, Lowe AM, Menon V, Slater JN, Webb JG, LeJemtel TH, Cotter G. Cardiac power is the strongest hemodynamic correlate of mortality in cardiogenic shock: a report from the SHOCK trial registry. J Am Coll Cardiol. 2004;44:340–348. doi: 10.1016/j.jacc.2004.03.060

9. Creager MA, Faxon DP, Halperin JL, Melidossian CD, McCabe CH, Schick EC, Ryan TJ. Determinants of clinical response and survival in patients with congestive heart failure treated with captopril. Am Heart J. 1982;104:1147–1154. doi: 10.1016/0002-8703(82)90043-6

10. Wilson JR, Schwartz JS, Sutton MS, Ferraro N, Horowitz LN, Reichek N, Josephson ME. Prognosis in severe heart failure: relation to hemodynamic measurements and ventricular ectopic activity. J Am Coll Cardiol. 1983;2:403–410. doi: 10.1016/s0735-1097(83)80265-4

11. Franciosa JA. Why patients with heart failure die: hemodynamic and functional determinants of survival. Circulation. 1987;75:IV20–27.

12. Lee WH, Packer M. Prognostic importance of serum sodium concentration and its modification by converting-enzyme inhibition in patients with severe chronic heart failure. Circulation. 1986;73:257–267. doi: 10.1161/01.cir.73.2.257

13. Opotowsky AR, Hess E, Maron BA, Brittain EL, Barón AE, Maddox TM, Alshawabkeh LI, Wertheim BM, Xu M, Assad TR, et al. Thermodilution vs Estimated Fick Cardiac Output Measurement in Clinical Practice: An Analysis of Mortality From the Veterans Affairs Clinical Assessment, Reporting, and Tracking (VA CART) Program and Vanderbilt University. JAMA Cardiol. 2017;2:1090–1099. doi: 10.1001/jamacardio.2017.2945

14. Unverferth DV, Magorien RD, Moeschberger ML, Baker PB, Fetters JK, Leier CV. Factors influencing the one-year mortality of dilated cardiomyopathy. Am J Cardiol. 1984;54:147–152. doi: 10.1016/0002-9149(84)90320-5

15. Garan AR, Kanwar M, Thayer KL, Whitehead E, Zweck E, Hernandez-Montfort J, Mahr C, Haywood JL, Harwani NM, Wencker D, et al. Complete Hemodynamic Profiling With Pulmonary Artery Catheters in Cardiogenic Shock Is Associated With Lower In-Hospital Mortality. JACC Heart Fail. 2020;8:903–913. doi: 10.1016/j.jchf.2020.08.012

16. Thayer KL, Zweck E, Ayouty M, Garan AR, Hernandez-Montfort J, Mahr C, Morine KJ, Newman S, Jorde L, Haywood JL, et al. Invasive Hemodynamic Assessment and Classification of In-Hospital Mortality Risk Among Patients With Cardiogenic Shock. Circ Heart Fail. 2020;13:e007099. doi: 10.1161/CIRCHEARTFAILURE.120.007099

17. Verbrugge FH, Tang WH, Mullens W. Renin-Angiotensin-aldosterone system activation during decongestion in acute heart failure: friend or foe? JACC Heart Fail. 2015;3:108–111. doi: 10.1016/j.jchf.2014.10.005

18. Heidenreich PA, Bozkurt B, Aguilar D, Allen LA, Byun JJ, Colvin MM, Deswal A, Drazner MH, Dunlay SM, Evers LR, et al. 2022 AHA/ACC/HFSA Guideline for the Management of Heart Failure: A Report of the American College of Cardiology/American Heart Association Joint Committee on Clinical Practice Guidelines. J Am Coll Cardiol. 2022;79:e263–e421. doi: 10.1016/j.jacc.2021.12.012

19. Shore S, Golbus JR, Aaronson KD, Nallamothu BK. Changes in the United States Adult Heart Allocation Policy: Challenges and Opportunities. Circ Cardiovasc Qual Outcomes. 2020;13:e005795. doi: 10.1161/CIRCOUTCOMES.119.005795

